# Intrinsic Susceptibility Of The Left Vs Right Recurrent Laryngeal Nerve To Paralysis: A Systematic Review

**DOI:** 10.1101/2023.08.09.23293889

**Authors:** Aaron Oswald, Michael J Pitman

## Abstract

**Objective:** Unilateral vocal fold paralysis (UVFP) occurs more commonly on the left than right, a difference historically attributed to greater left recurrent laryngeal nerve (RLN) length and its presence in the thorax with consequential increased exposure to injury. However, considering the importance of mRNA locally transcribed in the distal nerve after nerve injury, there may be other intrinsic neuromuscular reasons for this discrepancy. To investigate whether intrinsic neuromuscular issues influence laterality, this study investigates the rate of paralysis relative to side in idiopathic and short-term intubation cases, excluding cases due to identifiable disease and surgery.

**Data Sources:** Embase and PubMed database

**Review Methods:** A systematic literature search was performed to capture articles published up until May 2022. Articles were included if laterality and etiology of paralysis were reported. Demographic data was extracted for patients diagnosed with idiopathic paralysis or paralysis after intubation for procedures performed outside the head, neck, and thorax. Study design was collected from included articles.

**Results:** Twenty-one studies were included, from which 702 patients were drawn for analysis. Within the idiopathic group, 69.2% were left-sided. Within the post-intubation group, 67.9% were left-sided.

**Conclusions:** The available evidence indicates that left-sided paralysis is more common in patients with idiopathic or short-term intubation related UVFP. This suggests intrinsic neuromuscular differences contribute to the left RLN being more susceptible than the right to damage and dysfunction. Further study is needed to identify these differences, which may provide insights into the pathophysiology of RLN paralysis as well possible therapeutic options.

## Introduction

Unilateral vocal fold paralysis (UVFP) is commonly seen in otolaryngology practices, and leads to significant morbidity related to dysphagia and aspiration, hoarseness, difficulty with communication, and dyspnea.^1^ Much research has been devoted to understand the pathophysiology of RLN paralysis and developing therapeutics that improve treatment through functional reinnervation.^2-6^ One of the clues to improving reinnervation after injury may lie in the clinical appreciation of paralysis occurring more frequently on the left side. If there are inherent neuromuscular disparities in the processes of reinnervation on the left versus right after injury, these targets may be exploited therapeutically.

There are many known etiologies of UVFP, including systemic diseases, as well as disruption or injury along the recurrent laryngeal nerve, proximal vagus nerve, and motor pathways within the brain. Due to its increased length and course, the left recurrent laryngeal nerve is more susceptible to malignancy or iatrogenic injury, from thoracic malignancy, and cardiac, thoracic, or cervical surgery.^7^ In addition to these obvious reasons why the longer left RLN results in more frequent paralysis, it is also felt the length of the nerve itself makes reinnervation more difficult and results in more energy expended as proteins transcribed in reaction to injury must travel between the cell body and the nerve growth cone.^8-10^

Findings over the last 20 years suggest length alone may not be the reason for this laterality, as significant protein transcription after nerve injury takes place locally, in the distal nerve. M s are transported down the axon in advance and their translation is activated by injury or other stimuli. By transcribing mRNA locally, nerves can respond rapidly to injury, distinct from proteins in the cell body.^11^

Studies on rates of nerve recovery after injury also cast doubt on the importance of axon distance. One study on EMG recovery after injury in the canine larynx found no difference in rates of recovery between injuries 5 cm from the cricoid, and 5 cm further inferior.^12^ Another study modeling voice recovery after recurrent laryngeal nerve injury found that the extent of injury, rather than the location along the nerve, determined the kinetics of recovery.^13^

Studies have evidenced the importance of local translation after injury of peripheral axons.^14^ Netrin-1 and its receptors have also been shown to play significant roles in local translation.^15,16^ Accordingly, studies of RLN regeneration in the rat suggest that Netrin-1 may play a significant role in axon guidance and reinnervation.^2,17,18^

The present study aims to better elucidate the intrinsic susceptibility of the left recurrent laryngeal nerve to paralysis compared to the right nerve, by determining whether left-sided UVFP is more common in cases where only neuromuscular factors would influence paralysis. Our primary question was as follows: In a group of patients without a known anatomic factor predisposing them to recurrent laryngeal nerve paralysis on one side over the other, do the remaining etiologies for vocal fold paralysis lead to a predominance of paralysis on the left side over the right side? To this end, a systematic review was performed comparing the rate of paralysis relative to side in idiopathic and short-term intubation cases.

### Search Strategy

A search of the Embase and PubMed databases was conducted. The database was searched from inception until August 2022. Subject headings were used, and keywords included “vocal fold” and “vocal cord”; “paralysis”, “paresis, “hypomobility”, “immobility”, “palsy”, or similar words; and “left” or “right”. This search yielded 2099 articles.

### Study Selection

Duplicates were removed with a first pass to automatically capture articles with identical titles and author lists, and with a second pass to capture potential duplicates that were missed because of slight differences in capitalization or formatting. All were reviewed manually before being removed. After removal of duplicates, 2068 articles remained. Screening of articles according to inclusion criteria and full text review were performed by two reviewers (A.O., M.P.). Full text review was performed for all articles that ultimately fulfilled inclusion criteria, and was performed for some but not all articles during screening, as needed to determine inclusion. Unpublished studies and presentations, dissertations, and studies not in English or without full-text English translations available were not included. Articles that did not report primary data on patients with vocal fold paralysis were excluded. This included systematic reviews and analysis articles, basic science studies, veterinary and animal studies, and studies that included overlapping sets of patients. Studies were only included if they reported laterality of vocal fold paralysis. Case reports and case series including less than four patients with unilateral vocal fold paralysis were excluded.

Only studies that reported etiology of vocal fold paralysis were included. Given our goal of learning about the RLN’s intrinsic susceptibility to damage, we excluded cases of RLN paralysis that were likely to be related to an etiology influenced by the nerve’s path from the skull base to larynx. Such cases included paralysis following surgery or procedures at any site at or adjacent to the course of the RLN including of the thyroid, parathyroid, central neck, mediastinum, heart, or thorax. We also excluded cases of disease, tumors, radiation or radiofrequency ablation occurring at the skull base or in proximity to the course of the recurrent laryngeal nerve. Examples include chest, neck, or skull base trauma, sarcoidosis, tuberculosis, syphilis, pulmonary embolism, pneumonia, pneumoconiosis, and other pulmonary disease. We also excluded stroke or other localized CNS causes. Finally, we excluded cases where neuropathy was unlikely to be the cause of vocal fold hypomobility, including laryngeal and pharyngeal tumors, cricoarytenoid joint fixation, and surgical cases where that RLN was not at risk, but the patient remained intubated following a surgery.

Only studies that reported laterality of their patients were included. Studies that selected patients with only left, only right, or only bilateral vocal fold paralysis were excluded. Studies that did not report information on laterality stratified to etiologic group were excluded. Some studies reported sets of patients in which some patients fulfilled our etiologic inclusion criteria and some did not. If the study otherwise fulfilled our inclusion criteria, but had less than four patients that fulfilled the criteria, that study was excluded. Based on these criteria, 21 articles were included (Fig. 1).

**Figure 1:** PRISMA flowchart for study screening

### Data Extraction

Sets of patients fulfilling our inclusion criteria were drawn from the included studies. Based on the above criteria, 702 patients from 21 articles were included. No overlap in patients was found between studies, which for the most part were from different medical centers or different populations. Two studies included adult patient sets from the same academic medical center in Taiwan, however patients included in those studies were diagnosed and treated in non-overlapping time frames (2002-2006 for one, 2011-2015 for the other).^19,20^

## Results

Characteristics of the 21 included studies are shown in Table 1. The majority of included patients were diagnosed with idiopathic UVFP. Each article included eligible patient sets from only one etiologic group. There were 660 patients with idiopathic UVFP drawn from 16 studies, and 28 patients with post-intubation UVFP drawn from 2 studies. All included post-intubation UVFP patients were intubated for procedures that do not put the RLN at risk, and were extubated immediately following surgery.^21,22^ The remaining 3 studies contained smaller groups of patients that were eligible for analysis, including 6 patients with multiple system atrophy, 4 patients with vincristine-related neuropathy, and 4 patients with Charcot-Marie-Tooth Disease.^23,24,25^

**Table 1:**
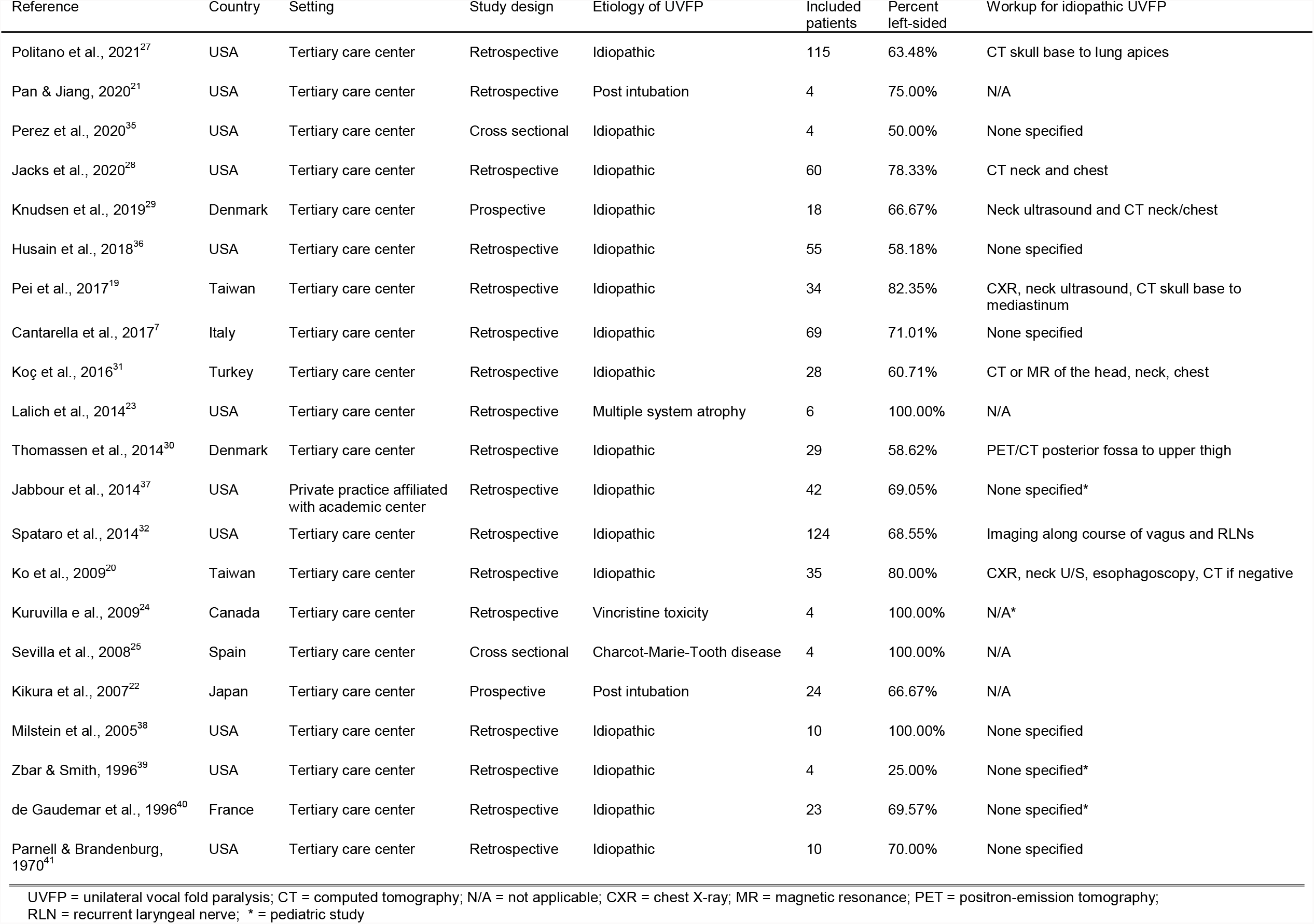
Characteristics of Included Studies.

| Reference | Country | Setting | Study design | Etiology of UVFP | Included patients | Percent left-sided | Workup for idiopathic UVFP |
| --- | --- | --- | --- | --- | --- | --- | --- |
| Politano et al., 2021<sup>27</sup> | USA | Tertiary care center | Retrospective | Idiopathic | 115 | 63.48% | CT skull base to lung apices |
| Pan & Jiang, 2020<sup>21</sup> | USA | Tertiary care center | Retrospective | Post intubation | 4 | 75.00% | N/A |
| Perez et al., 2020<sup>35</sup> | USA | Tertiary care center | Cross sectional | Idiopathic | 4 | 50.00% | None specified |
| Jacks et al., 2020<sup>28</sup> | USA | Tertiary care center | Retrospective | Idiopathic | 60 | 78.33% | CT neck and chest |
| Knudsen et al., 2019<sup>29</sup> | Denmark | Tertiary care center | Prospective | Idiopathic | 18 | 66.67% | Neck ultrasound and CT neck/chest |
| Husain et al., 2018<sup>36</sup> | USA | Tertiary care center | Retrospective | Idiopathic | 55 | 58.18% | None specified |
| Pei et al., 2017<sup>19</sup> | Taiwan | Tertiary care center | Retrospective | Idiopathic | 34 | 82.35% | CXR, neck ultrasound, CT skull base to mediastinum |
| Cantarella et al., 2017<sup>7</sup> | Italy | Tertiary care center | Retrospective | Idiopathic | 69 | 71.01% | None specified |
| Koç et al., 2016<sup>31</sup> | Turkey | Tertiary care center | Retrospective | Idiopathic | 28 | 60.71% | CT or MR of the head, neck, chest |
| Lalich et al., 2014<sup>23</sup> | USA | Tertiary care center | Retrospective | Multiple system atrophy | 6 | 100.00% | N/A |
| Thomassen et al., 2014<sup>30</sup> | Denmark | Tertiary care center | Retrospective | Idiopathic | 29 | 58.62% | PET/CT posterior fossa to upper thigh |
| Jabbour et al., 2014<sup>37</sup> | USA | Private practice affiliated with academic center | Retrospective | Idiopathic | 42 | 69.05% | None specified* |
| Spataro et al., 2014<sup>32</sup> | USA | Tertiary care center | Retrospective | Idiopathic | 124 | 68.55% | Imaging along course of vagus and RLNs |
| Ko et al., 2009<sup>20</sup> | Taiwan | Tertiary care center | Retrospective | Idiopathic | 35 | 80.00% | CXR, neck U/S, esophagoscopy, CT if negative |
| Kuruvilla et al., 2009<sup>24</sup> | Canada | Tertiary care center | Retrospective | Vincristine toxicity | 4 | 100.00% | N/A* |
| Sevilla et al., 2008<sup>25</sup> | Spain | Tertiary care center | Cross sectional | Charcot-Marie-Tooth disease | 4 | 100.00% | N/A |
| Kikura et al., 2007<sup>22</sup> | Japan | Tertiary care center | Prospective | Post intubation | 24 | 66.67% | N/A |
| Milstein et al., 2005<sup>38</sup> | USA | Tertiary care center | Retrospective | Idiopathic | 10 | 100.00% | None specified |
| Zbar & Smith, 1996<sup>39</sup> | USA | Tertiary care center | Retrospective | Idiopathic | 4 | 25.00% | None specified* |
| de Gaudemar et al., 1996<sup>40</sup> | France | Tertiary care center | Retrospective | Idiopathic | 23 | 69.57% | None specified* |
| Parnell & Brandenburg, 1970<sup>41</sup> | USA | Tertiary care center | Retrospective | Idiopathic | 10 | 70.00% | None specified |

UVFP = unilateral vocal fold paralysis; CT = computed tomography; N/A = not applicable; CXR = chest X-ray; MR = magnetic resonance; PET = positron-emission tomography; RLN = recurrent laryngeal nerve; \* = pediatric study

Risk of bias analysis for included studies is shown in Table 2. This was performed using the MINORS instrument.^26^

**Table 2:** Risk of Bias Assessment using the Methodological Index for Non-Randomized Studies (MINORS) Instrument.

| Reference | Clearly Stated Aim | Inclusion of Consecutive Patients | Prospective Collection of Data | Endpoints Appropriate to Study Aim | Unbiased Assessment of Study Endpoint | Follow-up Period Appropriate to Study Aim | Loss to Follow Up Less Than 5% | Prospective Calculation of Study Size | Adequate Control Group | Contemporary Groups | Baseline Equivalence | Adequate Statistical Analyses | Total Score | Risk of Bias |
| --- | --- | --- | --- | --- | --- | --- | --- | --- | --- | --- | --- | --- | --- | --- |
| Politano et al., 2021<sup>27</sup> | 2 | 2 | 0 | 2 | 1 | N/A | N/A | 0 | N/A | N/A | N/A | N/A | 7/16 | High |
| Pan & Jiang, 2020<sup>21</sup> | 2 | 1 | 0 | 2 | 0 | 2 | 1 | 0 | N/A | N/A | N/A | N/A | 8/16 | High |
| Perez et al., 2020<sup>35</sup> | 2 | 1 | 0 | 2 | 0 | N/A | N/A | 0 | N/A | N/A | N/A | N/A | 5/16 | High |
| Jacks et al., 2020<sup>28</sup> | 2 | 2 | 0 | 2 | 0 | N/A | N/A | 0 | N/A | N/A | N/A | N/A | 6/16 | High |
| Knudsen et al., 2019<sup>29</sup> | 2 | 2 | 2 | 2 | 0 | 2 | 1 | 0 | N/A | N/A | N/A | N/A | 11/16 | High |
| Husain et al., 2018<sup>36</sup> | 2 | 1 | 0 | 2 | 0 | 2 | 2 | 0 | N/A | N/A | N/A | N/A | 9/16 | High |
| Pei et al., 2017<sup>19</sup> | 2 | 2 | 0 | 2 | 0 | 2 | 2 | 0 | N/A | N/A | N/A | N/A | 10/16 | High |
| Cantarella et al., 2017<sup>7</sup> | 2 | 2 | 0 | 2 | 0 | N/A | N/A | 0 | N/A | N/A | N/A | N/A | 6/16 | High |
| Koç et al., 2016<sup>31</sup> | 1 | 2 | 0 | 2 | 0 | N/A | N/A | 0 | N/A | N/A | N/A | N/A | 5/16 | High |
| Lalich et al., 2014<sup>23</sup> | 2 | 2 | 0 | 2 | 0 | 2 | 1 | 0 | N/A | N/A | N/A | N/A | 9/16 | High |
| Thomassen et al., 2014<sup>30</sup> | 2 | 2 | 0 | 2 | 1 | N/A | N/A | 0 | N/A | N/A | N/A | N/A | 7/16 | High |
| Jabbour et al., 2014<sup>37</sup> | 1 | 2 | 0 | 2 | 0 | 2 | 1 | 0 | N/A | N/A | N/A | N/A | 8/16 | High |
| Spataro et al., 2014<sup>32</sup> | 2 | 2 | 0 | 1 | 0 | N/A | N/A | 0 | N/A | N/A | N/A | N/A | 5/16 | High |
| Ko et al., 2009<sup>20</sup> | 2 | 2 | 0 | 2 | 0 | 2 | 2 | 0 | N/A | N/A | N/A | N/A | 10/16 | High |
| Kuruville e al., 2009<sup>24</sup> | 2 | 1 | 0 | 1 | 0 | N/A | N/A | 0 | N/A | N/A | N/A | N/A | 4/16 | High |
| Sevilla et al., 2008<sup>25</sup> | 2 | 1 | 0 | 2 | 0 | N/A | N/A | 0 | N/A | N/A | N/A | N/A | 5/16 | High |
| Kikura et al., 2007<sup>22</sup> | 2 | 2 | 2 | 2 | 0 | 2 | 2 | 0 | N/A | N/A | N/A | N/A | 12/16 | High |
| Milstein et al., 2005<sup>38</sup> | 2 | 2 | 0 | 2 | 1 | 1 | 2 | 0 | N/A | N/A | N/A | N/A | 10/16 | High |
| Zbar & Smith, 1996<sup>39</sup> | 1 | 2 | 0 | 2 | 0 | 2 | 1 | 0 | N/A | N/A | N/A | N/A | 8/16 | High |
| de Gaudemar et al., 1996<sup>40</sup> | 1 | 2 | 0 | 1 | 0 | 2 | 1 | 0 | N/A | N/A | N/A | N/A | 7/16 | High |
| Parnell & Brandenburg, 1970<sup>41</sup> | 2 | 2 | 0 | 2 | 0 | N/A | N/A | 0 | N/A | N/A | N/A | N/A | 6/16 | High |

Scoring criteria: 0 (not reported), 1 (reported but inadequate), or 2 (reported and adequate)

Among the idiopathic UVFP group, 453/660 patients were left-sided (69.2%). In the post-intubation group, 19/28 were left-sided (67.9%). Combining those two groups yields a rate of left-sided paralysis of 68.6%. The frequency of left-sided paralysis ranged from 50% to 100%. Removing studies with less than 20 patients, the frequency of left-sided paralysis ranged from 58.2% to 82.4%. Of the 14 patients with the neuropathic disease, all had left-sided UVFP.

Diagnosis of idiopathic UVFP was determined by an absence of features on history and physical exam pointing to an etiology, as well a radiologic examination, which was varied. Three of the 16 studies with included idiopathic UVFP patients specified protocols involving a dedicated CT of the neck.^27,28,29^ In one study, all patients underwent PET-CT.^30^ Two studies used protocols that involved a chest X-ray and neck ultrasound as screening tools, followed up by a CT only if those studies were negative, or as needed.^19,20^ Two other studies reported some imaging was used to work up patients diagnosed with idiopathic UVFP; one of those specified that cross-sectional imaging of the head, neck, and chest.^31,32^ The remaining 8 studies with idiopathic UVFP patients did not specify the radiologic workup.

## Discussion

UVFP is a frequently encountered disorder in otolaryngology practice that accounts for significant morbidity, particularly from dysphagia and dysphonia, leading to risk of aspiration pneumonia and impaired communication. While there are static surgical procedures available to treat UVFP, consistent restoration of normal vocal fold function will require proper reinnervation. Here we demonstrate a consistently increased frequency of left-sided paralysis in etiologies of UVFP independent from anatomic factors that would be expected to put one nerve at risk over the other. Confirming an increased rate of left sided paralysis, in cases of idiopathic paralysis or that after short term intubation, suggests the left RLN is inherently more susceptible to nerve injury or is less able to effectively regenerate and reinnervate the larynx. While this has been attributed to the length of the nerve, considering the importance of local protein translation in the axon after nerve injury, as well as data showing that location of injury along the RLN does not impact the rate of recovery, length is likely not the sole contributor to this discrepancy.^8-13^ It is reasonable to conjecture there are other intrinsic neuromuscular differences between the left and right RLN and the intrinsic laryngeal muscles they target that would be responsible for the disparity in the laterality rates of paralysis. The reason for these differences should be explored and may provide insights into the pathophysiology of RLN paralysis as well possible therapeutic options.

Other proposed explanations for the increased rate of left-sided paralysis among patients diagnosed with idiopathic UVFP may fall into several categories. One possibility is that a significant portion of patients who carry this diagnosis have an occult anatomic lesion leading to compression or damage to the RLN in one or more locations. This may preferentially affect the left RLN, given its longer course passing through the mediastinum. This is unlikely as such a lesion would need to be missed during imaging workup, or be below the resolution of the imaging used.

A possible occult anatomic etiology proposed in recent literature is aortic arch compliance. Recent investigations have demonstrated an association between increased aortic arch compliance and idiopathic left-sided UVFP, when compared to normal matched controls. Aortic arch compliance was determined by measuring changes in aortic arch diameter in a cardiac-gated MRI.^33,34^ In those studies, each patient had been previously diagnosed with idiopathic UVFP after undergoing radiologic evaluation of the chest and neck prior.

Length-dependent mechanisms of neuropathy resulting in a higher rate of left sided paralysis have been proposed for Charcot-Marie-Tooth disease, and for vincristine-related neurotoxicity.^24,25^ In each of these groups, as well as the group with multiple system atrophy, all patients with UVFP were left-sided paralysis only, though the numbers of eligible patients were small.^23,24,25^ Vincristine-related neurotoxicity has been associated with shortened and decreased quantity of microtubules, which could put the longer axons of the left RLN at higher risk. The authors propose that the progression of neuropathy in a length-dependent manner makes an isolated right-sided paralysis less likely to occur.^24^ Increased rates of left-sided paralysis related to Charcot-Marie Tooth disease were also attributed to length-related neuropathy that occurs in other nerves in the disease.^25^ While it is known that patients with multiple system atrophy tend to have vocal fold abductor weakness, and many of them present with bilateral vocal fold paralysis, the authors did not have a disease-specific explanation for the preference for left-sided UVFP.^23^

It is interesting to note that the rate of left-sided paralysis is similar in post-intubation UVFP and idiopathic UVFP. The tendency of left-sided paralysis in the post-intubation setting has been postulated to be related to right-handed technique during intubation, or placement of the endotracheal tube at the right side of the mouth, angling the distal portion of the tube to the left,^21^ though this explanation would be difficult to verify. Regardless, compression from the cuff of an endotracheal tube against the subglottis should lead to relatively similar neurovascular compression on both RLNs, via direct compression or perineural edema. Even if there is some discrepancy in tube pressures, the extreme difference in laterality of post-intubation paralysis (67.9% on the left) suggests an increased intrinsic susceptibility to paralysis on the left.

Limitations of this review lie in the heterogeneity of studies and samples from which the patients were drawn. Although all patients were examined by otolaryngologists at academic or tertiary care centers, we were unable to verify the clinical factors and workup leading to a diagnosis of idiopathic UVFP in all cases.

## Conclusion

This systematic review of studies entailing 688 patients indicates that idiopathic and intubation-related UVFP preferentially affects the left side over the right side. This is likely influenced by intrinsic neuromuscular differences between the left and right RLN, beyond just nerve length. Further investigations are necessary to identify these differences which may provide insights into the pathophysiology of RLN paralysis as well possible therapeutic options.

## Data Availability

All data produced in the present study are available upon reasonable request to the authors
All data produced in the present work are contained in the manuscript

